# Mechanical Thrombectomy for Distal Medium Vessel Occlusions in Acute Ischemic Stroke: A Systematic review and Meta-Analysis of Randomized Controlled Trials

**DOI:** 10.1101/2025.06.26.25330382

**Authors:** Meena Chandu, Prachi Mohapatra, Pradeep Kumar, Rajesh Kumar Singh, Deepti Vibha

**Affiliations:** Department of Neurology, All India Institute of Medical Sciences, New Delhi, India; Clinical research unit, All India Institute of Medical Sciences, New Delhi, India

**Keywords:** Mechanical thrombectomy, acute ischemic stroke, distal medium vessel occlusion, endovascular therapy, functional outcome, randomized controlled trials

## Abstract

**Background:** Mechanical thrombectomy (MT) is established as standard care for acute ischemic stroke (AIS) due to large vessel occlusion, but its role in distal medium vessel occlusions (DMVOs) remains uncertain. We conducted a systematic review and meta-analysis to evaluate the efficacy and safety of MT vs best medical management (BMM) in patients with DMVO-related AIS.

**Methods:** A systematic search of PubMed, Embase, Scopus, and the Cochrane Library was conducted through February 2025 to identify randomized controlled trials (RCTs) comparing MT to BMM in patients with DMVO. The primary outcome was functional independence at 90 days (modified Rankin Scale [mRS] 0–2). Secondary outcomes included excellent outcome (mRS 0–1), symptomatic intracerebral haemorrhage (sICH), all-cause mortality, and serious adverse events. Pooled odds ratios (ORs) with 95% confidence intervals (CIs) were calculated using a random-effects model and heterogeneity was assessed with the I^2^ statistic.

**Results:** Three RCTs involving 1,222 patients were included; 601 received MT and 623 received BMM. MT was not associated with improved functional independence at 90 days (OR, 0.81; 95% CI, 0.49–1.32). The risk of sICH was higher in the MT group (OR, 2.33; 95% CI, 1.31– 4.13), while mortality rates were similar (OR, 1.33; 95% CI, 0.90–1.98). In a subgroup of 466 patients with M2 occlusions, MT showed no significant benefit over BMM (OR, 0.93; 95% CI, 0.72–1.19).

**Conclusions:** MT did not improve functional outcomes in patients with DMVO, including those with M2 occlusions, and was associated with a higher risk of sICH. Further trials are warranted to refine patient selection and procedural approaches in this population.

## Introduction

Mechanical thrombectomy (MT) has revolutionized the management of acute ischemic stroke (AIS) due to large vessel occlusion (LVO), with multiple landmark randomized controlled trials (RCTs) demonstrating its superiority over best medical therapy in improving functional outcomes.^1^

Encouraged by these outcomes, there is growing interest in extending MT to more distal intracranial occlusions—collectively referred to as medium vessel occlusions (MeVOs)— commonly referring to occlusions beyond the M1 or basilar artery segments, (though the definition is not uniform across studies).^2^ These occlusions account for 25% to 40% of AIS cases, yet they have not been well represented in previous thrombectomy trials.^3,4^

Although MeVO strokes often present with milder initial deficits than LVOs, outcomes are not uniformly benign.^5^ Approximately one-third of patients fail to achieve functional independence despite receiving intravenous thrombolysis (IVT), and mortality can approach 9%. Moreover, IVT demonstrates suboptimal recanalization rates in this population, typically below 50%, limiting its effectiveness as a definitive therapy.^6^

Evidence from retrospective cohorts, prospective registries, and post hoc analyses—most prominently from the HERMES meta-analysis—supports the feasibility and potential benefit of mechanical thrombectomy in anatomically and clinically selected MeVO cases, especially in dominant M2 occlusions, with high reperfusion rates and favourable functional outcomes.^7,8^ However, these findings are largely observational and derived from heterogenous cohorts, making it difficult to draw definitive conclusions.

Most MT trials underrepresented patients with MeVOs, and current guidelines reflect this evidence gap. The American Heart Association/American Stroke Association (AHA/ASA) provides a class IIb recommendation for MT in select M2 occlusions within 6 hours of onset,^9^ while the European Stroke Organisation (ESO) guidelines cautiously support its use in proximal MeVOs but refrain from endorsing intervention in more distal territories.^10^ These recommendations are largely based on observational data and expert consensus, underscoring the critical need for high-quality randomized evidence to guide practice in this growing subset of AIS. Notably, despite limited guideline endorsement, clinical interest and intervention in MeVO and distal vessel occlusion (DVO) stroke appear to be increasing, as reflected by the growing number of publications in recent years.^11^

To address this unmet need, several RCTs are now underway evaluating the efficacy and safety of MT in MeVO stroke. We conducted a systematic review and meta-analysis of randomized controlled trials (RCTs) to synthesize the current evidence on clinical outcomes, angiographic success, and procedural safety of MT for MeVOs.

## Methods

### Search Strategy and Study Selection

This systematic review and meta-analysis was conducted in accordance with PRISMA 2020 (Preferred Reporting Items for Systematic Reviews and Meta-Analyses) guidelines and was prospectively registered in the PROSPERO database (CRD420250651779). A comprehensive literature search was performed across PubMed, Embase, Scopus, and the Cochrane Library from inception through February 2025 to identify randomized controlled trials (RCTs) comparing mechanical thrombectomy (MT) to best medical management (BMM) in patients with acute ischemic stroke (AIS) due to distal or medium vessel occlusions (DMVOs/MeVOs).

The search strategy incorporated combinations of the following terms: *“acute ischemic stroke” OR “ischemic stroke” OR “cerebral infarction”* AND *“mechanical thrombectomy” OR “endovascular therapy” OR “EVT”* AND *“medium vessel occlusion” OR “distal vessel occlusion*.*”* Additional records were identified through manual screening of references. Two independent co-authors (MC and PM) conducted title and abstract screening, followed by full-text review to determine final eligibility using Rayyan. (https://www.rayyam.ai) Any discrepancy was resolved by an independent author (DV).

### Eligibility Criteria

Studies were included if they met the following criteria:

- Design: Randomized controlled trials
- Population: Adults (≥18 years) with AIS due to occlusion of medium or distal intracranial vessels (i.e., A1–A5 segments of the anterior cerebral artery, M2–M4 segments of the middle cerebral artery, and P1–P5 segments of the posterior cerebral artery)
- Intervention: Mechanical thrombectomy performed within 24 hours of symptom onset
- Comparator: Best medical management without thrombectomy
- Outcome: Functional outcome assessed by the modified Rankin Scale (mRS) at 90 days

Studies were excluded if they were observational (prospective or retrospective), case reports or series, reviews, or lacked 90-day mRS outcomes.

### Data Extraction and Outcomes

Two reviewers independently extracted data using a standardized form. The following details were extracted: study characteristics (first author, year, setting, sample size), baseline demographics (age, sex, comorbidities, National Institutes of Health Stroke Scale (NIHSS), Alberta Stroke Program Early CT Score (ASPECTS), prior IV thrombolysis), procedural details (device type, recanalization rate), and outcome measures. Any missing effect estimates were calculated from raw event data when available. No imputation was performed. All outcomes were converted to a uniform format (odds ratios with 95% confidence intervals) for synthesis.

Primary Efficacy Outcome:

- Functional dependence, defined as modified Rankin Scale (mRS) score of 0-2 at 90 days.

Secondary Efficacy Outcomes:

- Excellent functional outcome (mRS score of 0-1) at 90 days.
- Functional outcome in the M2 occlusion subgroup (as reported by included studies using pooled odds ratios)

Safety Outcomes:

- Symptomatic intracerebral haemorrhage (sICH)
- All-cause mortality at 90 days
- Serious adverse events (SAEs)

### Risk of Bias Assessment

Risk of bias was assessed using the Cochrane Risk of Bias 2.0 (ROB2) tool, evaluating domains such as randomization process, deviations from intended interventions, missing outcome data, measurement of outcomes, and selection of reported results. Discrepancies were resolved through consensus or third-party adjudication.

### Statistical Analysis

Data synthesis was conducted using Review Manager (RevMan) version 5.3 and R software (R Foundation for Statistical Computing), employing the meta and metafor packages. For all dichotomous outcomes—including functional independence, mortality, and symptomatic intracerebral haemorrhage (sICH)—pooled odds ratios (ORs) with 95% confidence intervals (CIs) were calculated using a random-effects model to account for inter-study heterogeneity. Study outcome data were systematically tabulated using standardized extraction forms. Forest plots were generated to visually display individual study estimates and pooled effects. Statistical heterogeneity was assessed using the I^2^ statistic, with I^2^ >50% considered indicative of substantial heterogeneity. A sensitivity analysis was performed by restricting the analysis to studies that used an anatomical definition of the M2 segment.

## Results

### Study Characteristics

Out of 3,665 records screened, three randomized controlled trials comprising 1,222 patients with distal or medium vessel occlusion (DMVO) were included in the meta-analysis comparing outcomes between endovascular thrombectomy (EVT) and best medical management (BMM) (**Figure 1**). The baseline characteristics of patients were clinically comparable across studies, with similar age, sex distribution, comorbidities, and stroke severity (median NIHSS 6–8). ESCAPE-MeVO included approximately 85% M2/M3 occlusions, with a small number of ACA and PCA cases, while DISTAL included around 70% M2/M3 and a larger proportion of M4, ACA, and PCA occlusions, reflecting greater anatomical heterogeneity. These differences may affect procedural complexity and interpretation of pooled outcomes.

**Figure 1:**
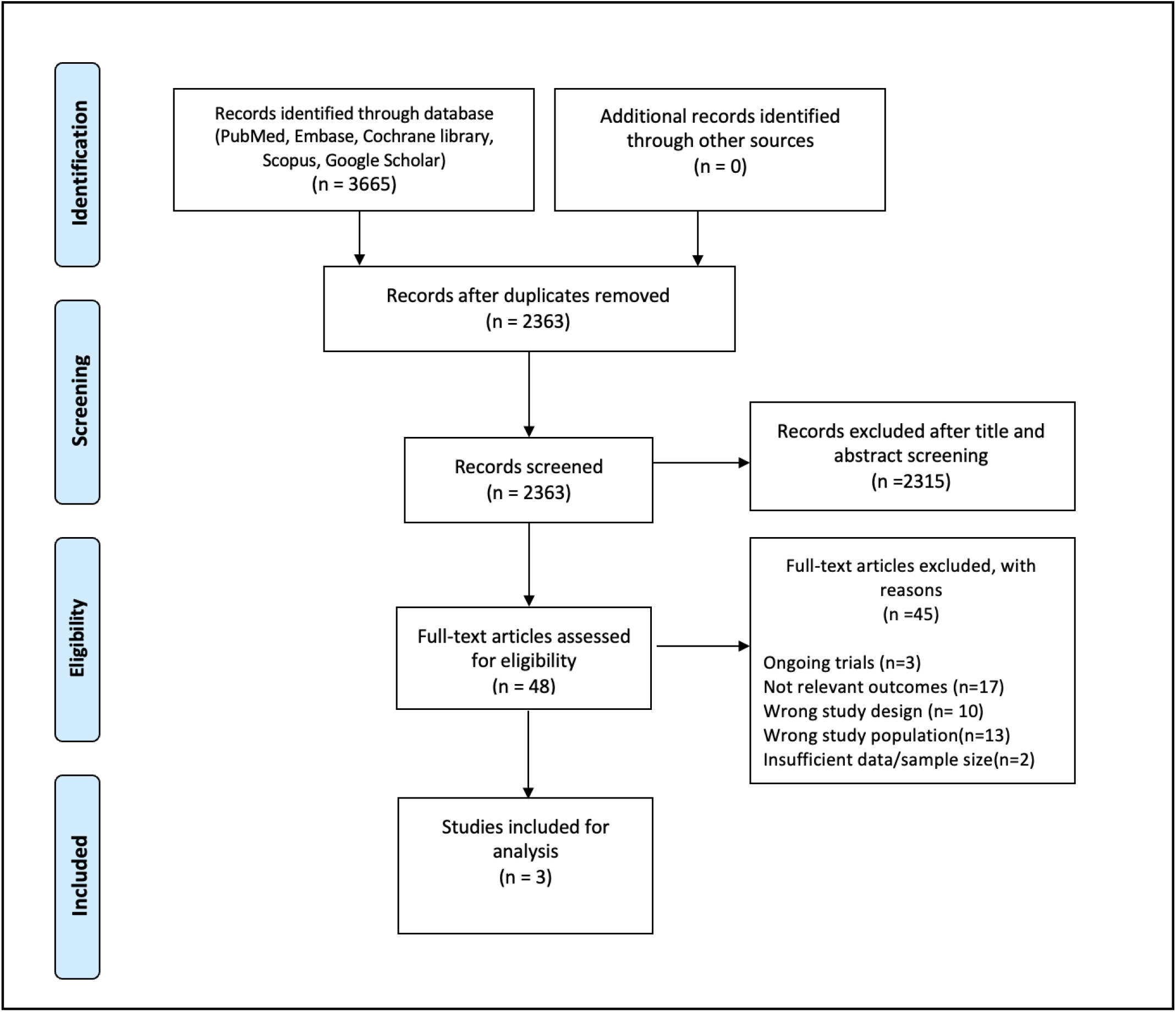
PRISMA flow chart

### Risk of bias assessment

The risk of bias was evaluated using the Cochrane Risk of Bias 2.0 tool across all included randomized controlled trials. Two studies were judged to have a low risk of bias across most domains, with the exception of performance bias due to their open-label (PROBE) design. While blinding of outcome assessors helps mitigate detection bias, it does not eliminate the potential influence of unblinded treatment delivery on patient behaviour or clinical management. One study (Clarençon et al.) was rated as having some concerns, primarily due to incomplete availability of trial data at the time of assessment, as full results have not yet been published. No study was judged to be at high risk of bias. A summary of domain-level risk assessments is presented in **Figure 2**.

**Figure 2:**
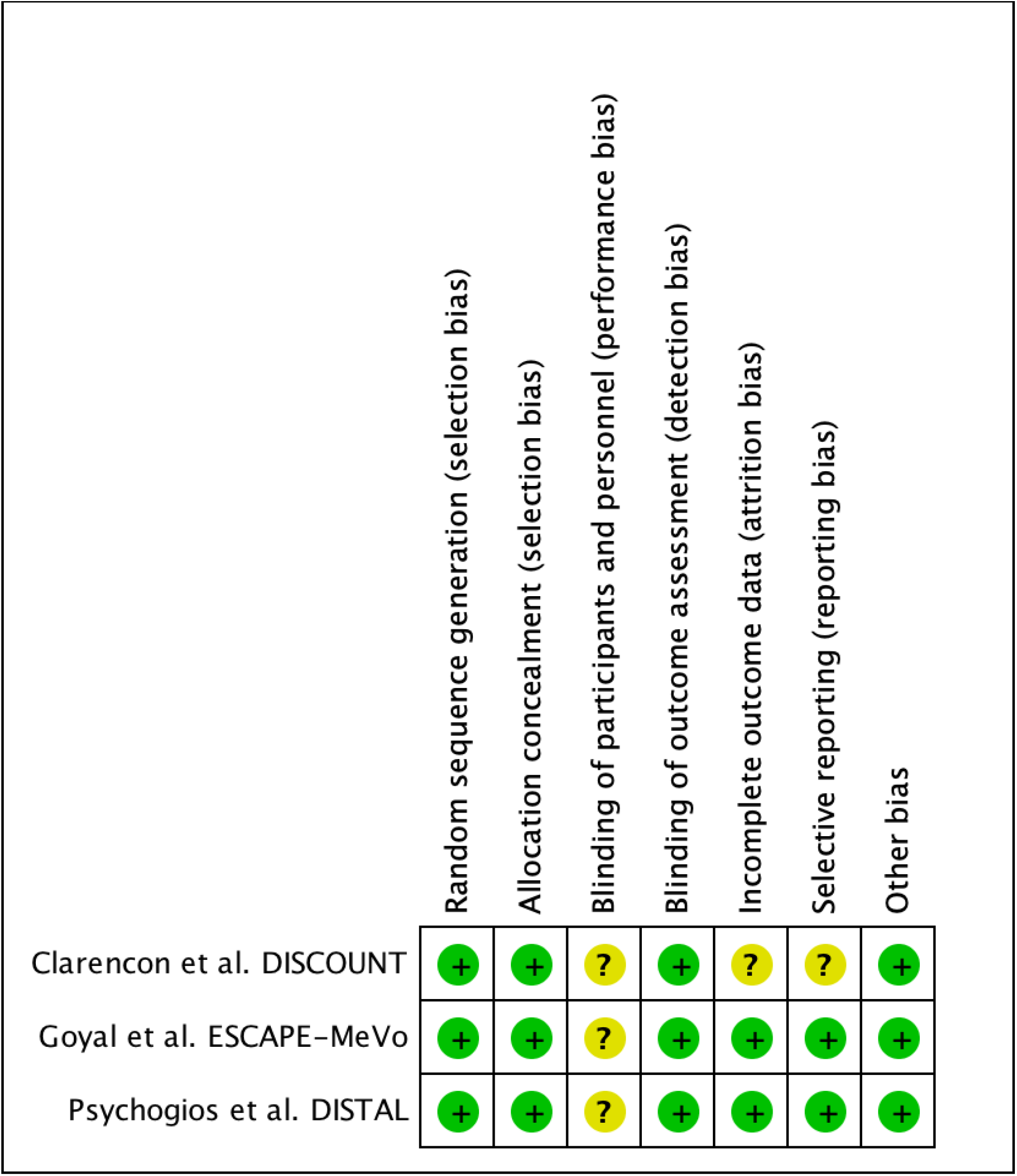
Risk of Bias Summary Across Domains for Included Randomized Trials (Cochrane Tool)

### Outcomes

#### Functional outcomes at 90 days

Three studies (Goyal et al., Psychogios et al., and Clarençon et al.) evaluated functional independence (modified Rankin Scale [mRS] 0–2) at 90 days. The pooled odds ratio (OR) was 0.81 (95% CI: 0.49–1.32), with moderate heterogeneity (I^2^ = 57.3%, p = 0.0961), indicating no significant benefit of endovascular therapy (EVT) over best medical management (BMM) (**Figure 3a**).

**Figure 3.**
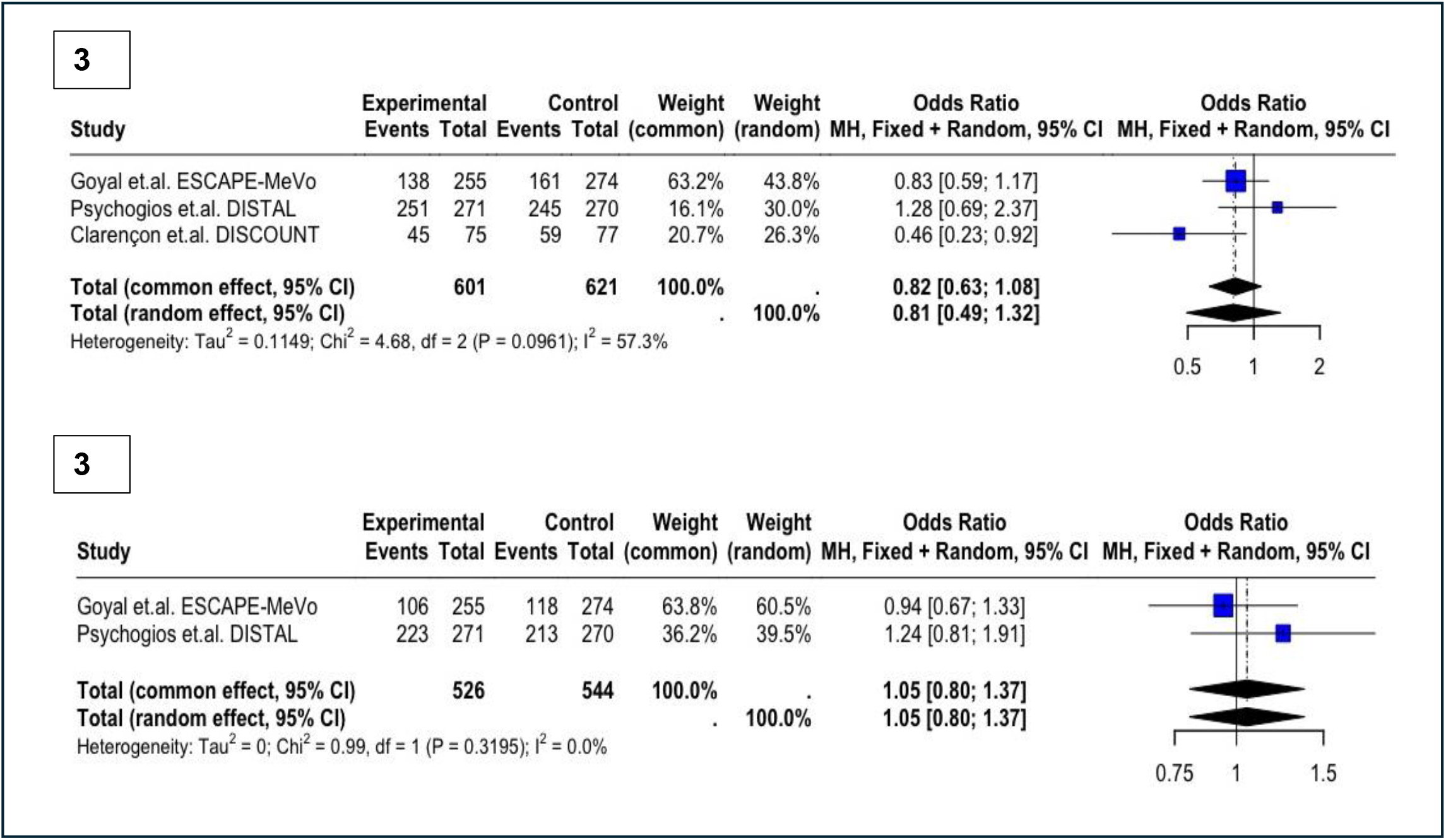
(A) Functional independence at 90 days (mRS 0-2) (B) Excellent functional outcome at 90 days (mRS 0–1) (EVT vs. BMM)

For excellent functional outcomes (mRS 0–1 at 90 days), two studies (Goyal et al. and Psychogios et al.) were included. The pooled OR was 1.05 (95% CI: 0.80–1.37), with no heterogeneity (I^2^ = 0%, p = 0.3195), again showing no significant difference between EVT and BMM (**Figure 3b**).

In a subgroup analysis of patients with M2 occlusions, data from three studies (Bejoy et al., Goyal et al., and Psychogios et al.) yielded a pooled OR of 0.96 (95% CI: 0.71–1.21), with low heterogeneity (I^2^ = 8.0%, p = 0.337), also demonstrating no significant difference between groups (**Figure 4**).

**Figure 4.**
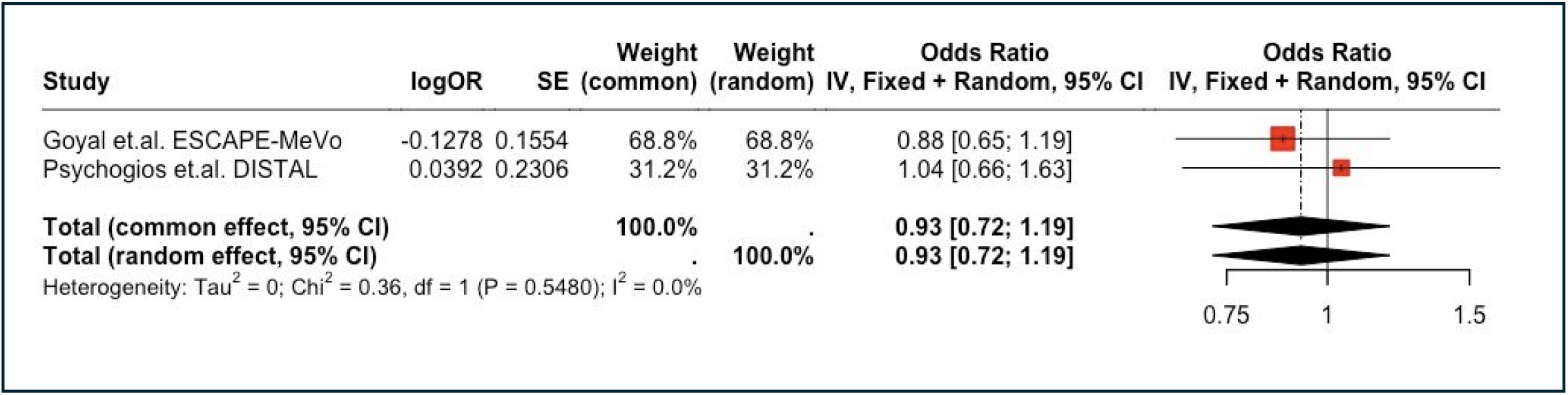
Functional Outcome in M2 Occlusion: EVT vs. BMM (Pooled Odds Ratio)

#### Symptomatic intracerebral haemorrhage (sICH)

Three studies (Goyal et al., Psychogios et al., and Clarençon et al.) assessed the incidence of sICH. The pooled OR was 2.33 (95% CI: 1.31–4.13), with no observed heterogeneity (I^2^ = 0%, p = 0.9353), indicating a higher rate of sICH in the EVT group compared to BMM (**Figure 5**).

**Figure 5.**
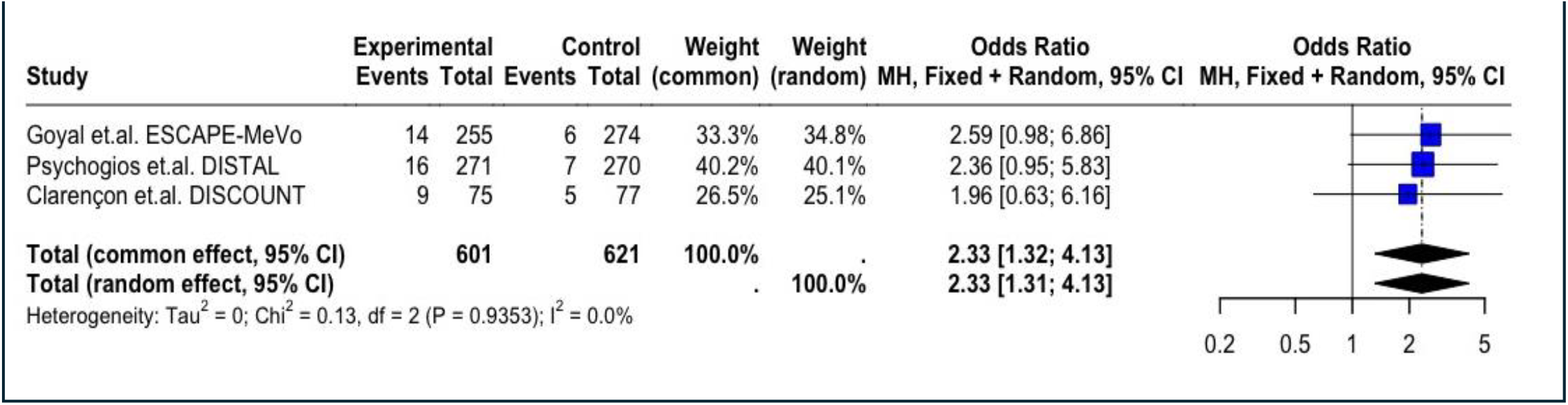
Symptomatic intracerebral haemorrhage (sICH): EVT vs. BMM

#### All-cause mortality at 90 days

Two studies (Goyal et al. and Psychogios et al.) reported data on all-cause mortality. The pooled OR was 1.33 (95% CI: 0.90–1.98), with low heterogeneity (I^2^ = 14.5%, p = 0.2793), suggesting no statistically significant difference between EVT and BMM (**Figure 6a**).

**Figure 6.**
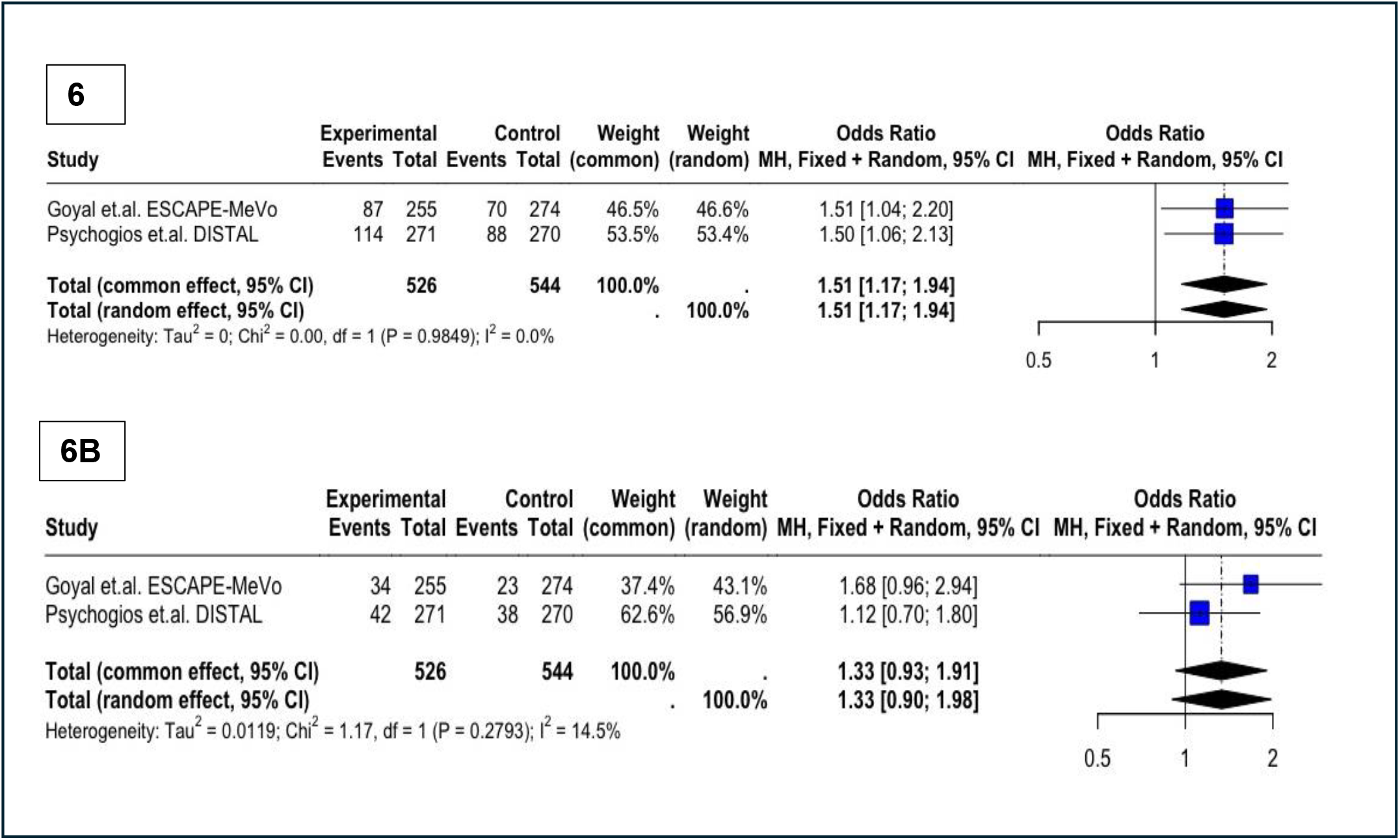
(A) All-cause mortality at 90 days (B) Serious adverse events(EVT vs. BMM)

#### Serious adverse events (SAEs)

Two studies (Goyal et al. and Psychogios et al.) evaluated the occurrence of SAEs. The pooled OR was 1.51 (95% CI: 1.17–1.94), with no heterogeneity (I^2^ = 0%, p = 0.9849), indicating a higher incidence of SAEs in the EVT group compared to the BMM group (**Figure 6b**) (outcomes summarised in **Table 1**)

**Table 1.**
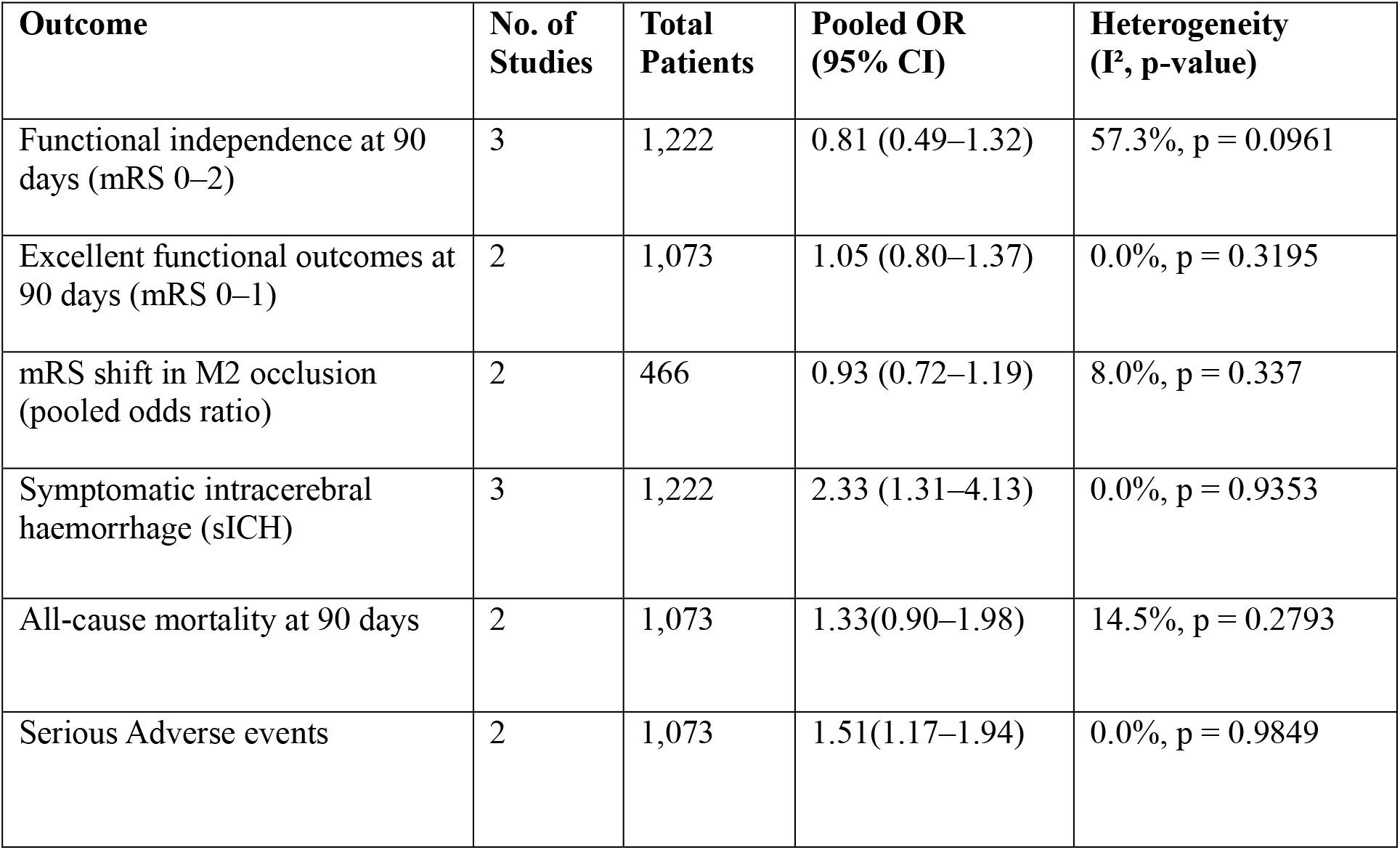
Summary of efficacy and safety outcomes in DMVO stroke.

## Discussion

This meta-analysis summarizes evidence from randomized controlled trials comparing endovascular thrombectomy (EVT) with best medical management (BMM) in patients with AIS due to distal medium vessel occlusion (DMVO) presenting within 24 hours. The primary outcome, functional independence at 90 days (mRS 0–2), did not significantly differ between EVT and BMM groups. Similarly, no difference was observed in excellent functional outcomes (mRS 0–1 at 90 days). Several factors may explain why our meta-analysis did not demonstrate a consistent benefit of EVT in DMVO stroke:

Firstly, reported recanalization rates in DMVO trials have been modest—ranging from 71% to 75%—despite the use of the MeVO-eTICI classification, in contrast to the 80–90% success rates typically achieved in large vessel occlusion (LVO) thrombectomy trials.^12–14^ In the ESCAPE-MeVO and DISTAL trials, stent retrievers were used as the first-line treatment. Although stent retrievers are highly effective for LVOs, they may carry increased procedural risks under deployment and removal in smaller, more tortuous arteries, such as those involved in MeVOs and DVOs. This highlights the technical challenges of achieving effective reperfusion in smaller-caliber distal vessels.

Secondly, the overall time from symptom onset to recanalization was substantially longer in DMVO trials. For instance, the ESCAPE-MeVO trial reported a median recanalization time of 359 minutes compared to 241 minutes in the original ESCAPE trial. These delays may be due to a higher proportion of patients undergoing general anaesthesia (41% in ESCAPE-MeVO vs. 9% in ESCAPE), greater difficulty in detecting DMVOs on imaging—as reflected by longer imaging-to-puncture times in the DISTAL trial (70 vs. 60 minutes)—and the technical complexity of accessing distal vessels, which requires considerable operator expertise.^15,16^

Thirdly, the anatomical nature of DMVOs poses intrinsic limitations. These vessels are often end arteries with limited or absent collateral flow, reducing the volume of salvageable tissue and potentially attenuating the clinical benefit of reperfusion.

Fourth, patient selection may have influenced trial outcomes. A substantial proportion of patients had low baseline NIHSS scores (e.g., 41% with NIHSS <5 in the DISTAL trial) and advanced age. This raises the possibility that randomization was affected by physician discretion, particularly in patients perceived as either too mild or too frail for intervention. The lack of screening logs in several trials further limits our ability to assess selection bias and external validity.

Fifth, another possible reason for the neutral findings is the use of the modified Rankin Scale (mRS) as the primary outcome measure. Although widely accepted, the mRS may lack sensitivity to detect subtle or early neurological improvements, especially in patients with mild strokes. Incorporating alternative outcome measures, such as early NIHSS change or patient-reported quality-of-life metrics, may better capture clinically meaningful benefit in this population.

In the subgroup analysis of M2 occlusions, our meta-analysis also found no statistically significant benefit of EVT over BMM. This contrasts with earlier findings from the HERMES collaboration,^7^ which pooled individual patient data and suggested potential benefit in carefully selected M2 occlusions—particularly those involving proximal or dominant branches. However, the HERMES analysis included only 130 M2 cases, limiting its statistical power. A separate meta-analysis of 12 non-randomized studies reported favourable outcomes with EVT in M2 occlusions, though these findings are subject to selection bias and methodological limitations inherent to observational data.^8^ Importantly, the trials included in our analysis enrolled a heterogeneous mix of M2 occlusion types—proximal, distal, dominant, and nondominant. Given the likely underrepresentation of proximal, dominant M2 occlusions (excluded in DISTAL trial) these findings should be interpreted with caution, as they may better reflect outcomes in non-dominant or distal M2 subtypes.

In terms of safety outcomes, our meta-analysis identified a higher incidence of symptomatic intracerebral haemorrhage (sICH) in patients treated with EVT compared to those receiving BMM. Although the absolute rate of sICH remained low, this consistent finding across trials raises important concerns regarding procedural safety in smaller, more fragile distal vessels. Despite the increased sICH risk, all-cause mortality did not significantly differ between groups, suggesting that sICH was not the primary driver of mortality. These findings underscore the need for further refinement of thrombectomy techniques and devices tailored to the anatomical and technical challenges of DMVO.

In light of these considerations, identifying which patients may benefit from EVT in the context of DMVO remains a central challenge. Ongoing trials—including FRONTIER-AP (ACTRN12621001746820p) and DISTALS (NCT 05152524) —are expected to refine our understanding of occlusion-specific efficacy, optimize patient selection strategies, and evaluate device performance tailored to distal occlusions.

A notable strength of this meta-analysis is its exclusive inclusion of randomized controlled trials and systematic evaluation of both efficacy and safety endpoints. Furthermore, its findings are supported by the early termination of the DISCOUNT trial ^17^ due to futility, reinforcing the conclusion that EVT, as currently practiced, may not significantly improve 90-day outcomes in DMVO stroke and carries a risk of procedural complications such as sICH. Nonetheless, several limitations should be acknowledged, including the small number of available trials, heterogeneity in DMVO definitions, variability in procedural techniques, and limited generalizability beyond M2 occlusions due to the small number of patients with more distal vessel involvement.

Although evidence from randomized trials remains inconclusive, individual patient-level data from HERMES and findings from observational studies suggest that EVT may benefit carefully selected patients with M2 occlusions. These insights highlight the need for individualized decision-making that considers clinical presentation, occlusion site, imaging characteristics, and procedural risk. Future research should aim to refine selection criteria using advanced imaging, establish standardized definitions for DMVO, and design well-powered trials focusing specifically on medium and distal occlusions. Additionally, the use of more sensitive functional outcome measures, shorter imaging-to-treatment times, and newer-generation thrombectomy devices may help define a more precise and effective role for EVT in DMVO stroke care.

## Data Availability

Available on request

## Abbreviations

AIS: acute ischemic stroke
BMM: best medical management
CI: confidence interval
DMVO: distal medium vessel occlusion
EVT: endovascular thrombectomy
mRS: modified Rankin Scale
NIHSS: National Institutes of Health Stroke Scale
sICH: symptomatic intracranial haemorrhage
TICI: Thrombolysis in Cerebral Infarction.

## Acknowledgments

The authors thank all investigators and trial teams of the included studies for making their data publicly available.

